# Accelerometer-derived circadian rhythm and colorectal cancer risk in UK Biobank: a prospective cohort study

**DOI:** 10.64898/2026.04.03.26350124

**Authors:** Maung Ni Chan Chin (Chengqin Ni), Javier Alejandro Berrio

**Author notes:** Correspondence: Javier Alejandro Berrio PhD, Program of Occupational Safety and Health, Fundación Universitaria Tecnológico Comfenalco, Cartagena, Bolívar, 130001, Colombia. Presentations: A portion of this work has been accepted for presentation at the “Belt and Road” Initiative Teaching Support Project Meeting 2026; University of Yangon, Yangon, Myanmar.

## Abstract

**Background:** While total physical activity is a recognized modifier of cancer risk, accelerometer-derived digital phenotyping enables high-resolution mapping of circadian behavior. Whether these multidimensional patterns comprising step counts, sleep, physical activity, circadian rhythmicity, and light exposure independently influence the risk of incident colorectal cancer (CRC) has not been comprehensively evaluated

**Methods:** We performed an exposure-wide association study (ExWAS) of 224 accelerometer-derived metrics among 95,050 UK Biobank participants who were free of CRC at accelerometry. To comprehensively define circadian rhythm patterns, we systematically categorized these metrics into five core behavioral domains: step counts, sleep architecture, physical activity bouts, circadian rhythmicity, and light exposure. Hazard ratios (HRs) and 95% confidence intervals were estimated using Cox proportional hazards models with age as the underlying timescale.

**Results:** During a median follow-up of 8.5 years, 775 participants developed CRC (503 colon; 269 rectal). In minimally adjusted models, 121 metrics showed nominal significance (31 for overall CRC, 89 for colon, and 1 for rectal cancer). Protective associations were predominantly observed for metrics characterizing activity intensity and bout structure; notably, higher mean acceleration during 5-10 minute bouts of moderate-to-vigorous physical activity was associated with reduced CRC risk (HR 0.88 per SD). In contrast, no metrics within the defined sleep or light exposure domains reached nominal significance. These associations attenuated substantially following progressive adjustment for lifestyle and metabolic covariates, suggesting potential confounding or shared biological pathways.

**Conclusions:** Our findings identified specific behavioral phenotypes within a multidimensional framework of circadian rhythm, including step counts, physical activity intensity, and bout structure, as being associated with CRC risk. However, the marked attenuation of signals after multivariable adjustment suggests these markers may not serve as independent predictors. These results underscore the complexity of multidimensional circadian digital biomarkers and necessitate independent replication to clarify their utility in cancer risk stratification.

## Introduction

Colorectal cancer (CRC) remains a major source of cancer burden and is the second leading cause of cancer mortality in the United States, making prevention through modifiable behavioural factors a major public health priority^1^. Physical activity is among the most consistently cited candidate protective factors, but most of the evidence base relies on self-report and therefore emphasizes total volume rather than fine-grained behavioural structure^2, 3^. Wrist accelerometry offers a different view. Beyond overall activity volume, accelerometer data can quantify how movement is accumulated, how often sedentary time is interrupted, how activity is distributed across the 24-hour cycle and how strongly behaviour aligns with circadian organization^4, 5^. Device-based cohort analyses have also shown that wearable-derived activity captures meaningful prospective variation in future health risk, supporting its use for large-scale epidemiologic discovery^6^.

Those distinctions may matter for CRC. Experimental and observational work implicates insulin regulation, inflammation, immune surveillance and circadian biology in colorectal carcinogenesis^7, 8^. If so, behavioural patterning may be informative in addition to total activity. At the same time, many accelerometer-derived metrics are highly correlated, which creates a large and partially redundant hypothesis space. Recent multidimensional accelerometer work has suggested that circadian organization may be better represented by joint profiles of rest-activity rhythm, daytime activity, sleep and chronotype rather than by any single marker alone^9^. Colon and rectal cancers may also differ in their relationship to accelerometer-derived behaviours because these subsites differ in anatomy, exposure to luminal contents and molecular characteristics^10^.

UK Biobank provides an opportunity to explore these questions at scale. The accelerometer sub study includes week-long wrist-device measurements together with linked cancer outcomes and baseline covariates. We therefore performed a hypothesis-generating exposure-wide analysis of accelerometer-derived metrics and incident CRC outcomes. Our aim was not to establish a short list of definitive causal markers, but to map the nominal signal burden, evaluate how strongly it depends on model specification and identify candidate behavioural domains for replication.

## Methods

### Study population

We analyzed data from the UK Biobank, a large-scale prospective cohort of over 500,000 individuals aged 40 to 69 years recruited across the United Kingdom between 2006 and 2010^11^. Between 2013 and 2015, a subset of participants underwent seven day wrist-worn accelerometry. Our analytic population was restricted to participants with valid accelerometer data and no prevalent CRC at the time of accelerometry.

### Accelerometer processing

Participants wore a triaxial Axivity AX3 accelerometer on their dominant wrist continuously for seven days. Raw acceleration files, sampled at 100 Hz, were processed using the GGIR (v3.3-4) open-source software in R^12, 13^. Signal derivation employed 5-second epochs using the Euclidean Norm Minus One (ENMO) metric to isolate human movement from gravitational acceleration^12^. Sensor autocalibration was implemented to ensure a calibration error threshold below 0.01 g^12^. Non-wear time was identified via the 2023 GGIR algorithm using axis-specific variability and a 150-mg range threshold^12^; recordings with <16 valid hours per 24-hour window were excluded^13^.

### Multidimensional Circadian Phenotyping

To systematically define the circadian rhythm beyond total activity volume, we characterized 224 candidate metrics categorized into five core behavioral and environmental domains:

### Objective Measured Light Exposure

As the primary zeitgeber^14^ (time-giver) for the master circadian clock, accelerometer-measured ambient light exposure was quantified using a validated wrist-light processing framework^15–18^. Raw light measurement recorded at 100 Hz was extracted, calibrated, standardized and transformed to ensure cross-device consistency^15^. We derived 24-hour rhythmic profiles by aggregating these data into 48 half-hour bins per period, with values averaged across all valid days to represent a stable individual-level exposure^15^. To characterize the diurnal light-exposure landscape, we collapsed these bins into five contiguous dayparts: nighttime (bins 1–16), morning (17–24), afternoon (25–32), evening (33–40), and late-night (41–48)^19^. Total daily light was calculated as the mean across the full 48-bin period, representing the cumulative environmental input to the circadian system^15^.

### Step Counts

Step-count metrics were generated via the Verisense step algorithm (v2) integrated within the GGIR processing workflow^20, 21^. This validated algorithm was applied to 15-Hz resampled triaxial acceleration data to capture the rhythmic mechanical loading associated with human locomotion^12^. We derived median daily step counts as a measure of overall ambulatory volume. To characterize maximal sustained walking intensity, we quantified mean peak cadence over 1-, 5-, 10-, and 30-minute windows; notably, these metrics represent the highest observed stepping rates across the 24-hour period and do not necessary to be consecutive^22^. Furthermore, steps were functionally categorized based on their behavioral context: incidental steps, purposeful steps and intensity-specific accumulations including light physical activity (LPA) steps and moderate-to-vigorous physical activity (MVPA) steps^22^.

### Sleep

The Sleep-Period-Time (SPT) method was used to infer the sleep window, effectively defining the rest phase of the 24-hour cycle and its temporal alignment with the biological night. To provide a high-resolution characterization of sleep consolidation and circadian phase, we derived a comprehensive suite of digital phenotypes encompassing total sleep duration (TSD), sleep efficiency (SE), and markers of sleep fragility such as the Number of Nighttime Awakenings (NNA) and wake after sleep onset (WASO)^9, 12^. Furthermore, we quantified the precise timing of sleep onset, wake-up time, and mid-sleep time to capture the multidimensional integrity of the sleep-wake cycle and its synchronization with the endogenous circadian clock.

### Physical Activity

Movement was systematically categorized using established acceleration thresholds for light (LPA: 40–100 mg), moderate (100–400 mg), and vigorous (>400 mg) activity^12, 13^. Beyond overall acceleration volume, we implemented an 80% bout criterion to characterize the temporal accumulation of activity across multiple durations (1, 5, and 10 minutes), reflecting the sustained nature of behavioral episodes^13^. To capture the fine-grained structure and complexity of daytime behavior, we quantified behavioral fragmentation through an extensive set of metrics, including transition probabilities between active and sedentary states and the Gini index of bout distributions^23–27^. These digital phenotypes provide a high-resolution map of how individuals consolidate or fragment their movement, offering a proxy for the robustness of daytime physical behavior within the 24-hour cycle.

### Circadian Rhythmicity and Fragmentation

We quantified the robustness and temporal stability of the 24-hour rest-activity cycle using a complementary suite of parametric and non-parametric approaches. Central to this characterization, the Relative Amplitude (RA) was derived to evaluate the overall strength and consolidation of the circadian rhythm^28^. This was calculated by integrating the Most-active 10-hour (M10) and Least-active 5-hour (L5) acceleration levels across the entire record, providing a normalized measure of the contrast between peak daytime activity and nocturnal rest. To assess rhythmic synchronization, Interdaily Stability (IS) was employed to measure the alignment of behavioral patterns with the 24-hour solar cycle, while Intradaily Variability (IV) quantified the hourly fragmentation of the rest-activity sequence^25, 29, 30^. The intrinsic complexity and long-range correlation structure of the acceleration signal were evaluated via Detrended Fluctuation Analysis (DFA) to derive the Self-Similarity Parameter (SSP) and Activity Balance Index (ABI)^25, 31^. Furthermore, internal biological timing was rigorously defined using cosinor^32^ and extended-cosinor models, yielding estimates of the midline-estimating statistic of rhythm (Mesor), amplitude, and acrophase (phi). Finally, the first-order autocorrelation of multi-day activity (Phi) ^25,33^ was calculated to capture the multidimensional integrity of the endogenous circadian system.

### Outcome ascertainment

Incident colorectal cancer (CRC) cases were identified through linkage with NHS England and National Records of Scotland, NHS Central Register using International Classification of Diseases (ICD-10) codes C18–C20^34^. We evaluated three primary outcomes: total CRC (C18–C20), colon cancer (C18), and rectal cancer (C19–C20). All prevalent CRC diagnosed prior to the date of accelerometry were excluded.

### Covariates and statistical analysis

We conducted a hypothesis-generating exposure-wide association study (ExWAS), modeling each of the 224 accelerometer metrics individually against total CRC, colon, and rectal cancer using Cox proportional hazards regression. The Minimal Discovery Model adjusted for core confounders: sex, race, Townsend Deprivation Index, smoking status, alcohol consumption, season of wear, and family history of CRC. Missing covariate data were handled via categorical indicators or median imputation^35^.

To evaluate the robustness and independence of the observed signals, we implemented pre-specified Sensitivity Tiers: (1) an Adiposity/Metabolic tier (adding BMI and diabetes status); (2) an Independent Pattern tier (adjusting for total acceleration to isolate structural behavioral effects); and (3) domain-specific tiers adjusting for shift-work history or device-quality metrics.^35^

Given the exploratory objective, we prioritized nominal associations (two-sided P < 0.05) as a prioritization screen rather than confirmatory evidence. Temporal robustness was assessed via lag-time analyses, excluding the first one and two years of follow-up to mitigate reverse causation. Proportional hazards assumptions were verified using Schoenfeld residuals. Additionally, E-values^36^ were calculated to quantify potential sensitivity to unmeasured confounding. Interaction-first subgroup analyses were performed by sex and age (early onset <65 vs. late onset ≥65 years). All analyses were conducted in R version 4.5.2^37^.

## Results

### Study population

Following the exclusion of prevalent CRC and suboptimal accelerometry, the final analytic cohort comprised 95,050 participants, encompassing 776,456 person-years of observation over a median follow-up of 8.5 years. During this period, we identified 775 incident CRC cases, of which 503 were localized to the colon and 269 were rectal or rectosigmoid cancers.

Compared with non-cases, individuals who developed incident CRC were older at the time of accelerometer wear (mean 66.0 vs. 62.3 years), predominantly male (53.2% vs. 43.6%), and exhibited a higher metabolic risk profile, including elevated body mass index (mean 27.4 vs. 26.7 kg/m²) and a lower prevalence of never-smokers (48.8% vs. 57.3%). Notably, cases displayed lower overall physical activity volume, as evidenced by reduced mean acceleration (27.8 vs. 29.4 mg), and were more likely to have a family history of CRC (17.5% vs. 11.9%) (Table 1).

**Table 1.** Baseline characteristics by colorectal cancer status. Baseline characteristics of the 95,050 participants included in analysis.

| Characteristic | Non-CRC | CRC |
| --- | --- | --- |
| N | 94275 | 775 |
| Age, years | 62.3 (7.8) | 66.0 (6.8) |
| Sex, n (%) |  |  |
| Male | 41066 (43.6) | 412 (53.2) |
| Female | 53209 (56.4) | 363 (46.8) |
| Race, n (%) |  |  |
| White | 91045 (96.6) | 759 (97.9) |
| Non-White | 3230 (3.4) | 16 (2.1) |
| Townsend Deprivation Index | -1.7 (2.8) | -1.8 (2.9) |
| BMI, kg/m<sup>2</sup> | 26.7 (4.5) | 27.4 (4.7) |
| Smoking status, n (%) |  |  |
| Never | 54057 (57.3) | 378 (48.8) |
| Past | 33702 (35.7) | 339 (43.7) |
| Current | 6516 (6.9) | 58 (7.5) |
| Education, n (%) |  |  |
| Pre College | 27437 (29.1) | 244 (31.5) |
| Post College | 66838 (70.9) | 531 (68.5) |
| Alcohol status, n (%) |  |  |
| Never | 2733 (2.9) | 22 (2.8) |
| Past | 2587 (2.7) | 25 (3.2) |
| Current | 88955 (94.4) | 728 (93.9) |
| Total acceleration, mg | 29.4 (8.4) | 27.8 (7.9) |
| Shift work history, n (%) |  |  |
| Yes | 7656 (8.1) | 51 (6.6) |
| CRC family history, n (%) |  |  |
| Yes | 11216 (11.9) | 136 (17.5) |

### Primary exposure-wide screen

Across the 224 candidate accelerometer-derived exposures, the primary Minimal Discovery Model identified a landscape of 121 nominal associations (p<0.05) across the three clinical outcomes. This signal burden encompassed 31 associations for overall CRC, 89 for colon cancer, and a single association for rectal cancer (Table 2). The prioritized lead set was predominantly concentrated within domains characterizing activity intensity, bout frequency, and behavioral fragmentation. Notably, despite the multidimensional nature of our circadian framework, no metrics within the sleep architecture or environmental light exposure domains reached nominal significance in the primary discovery model.

**Table 2.**
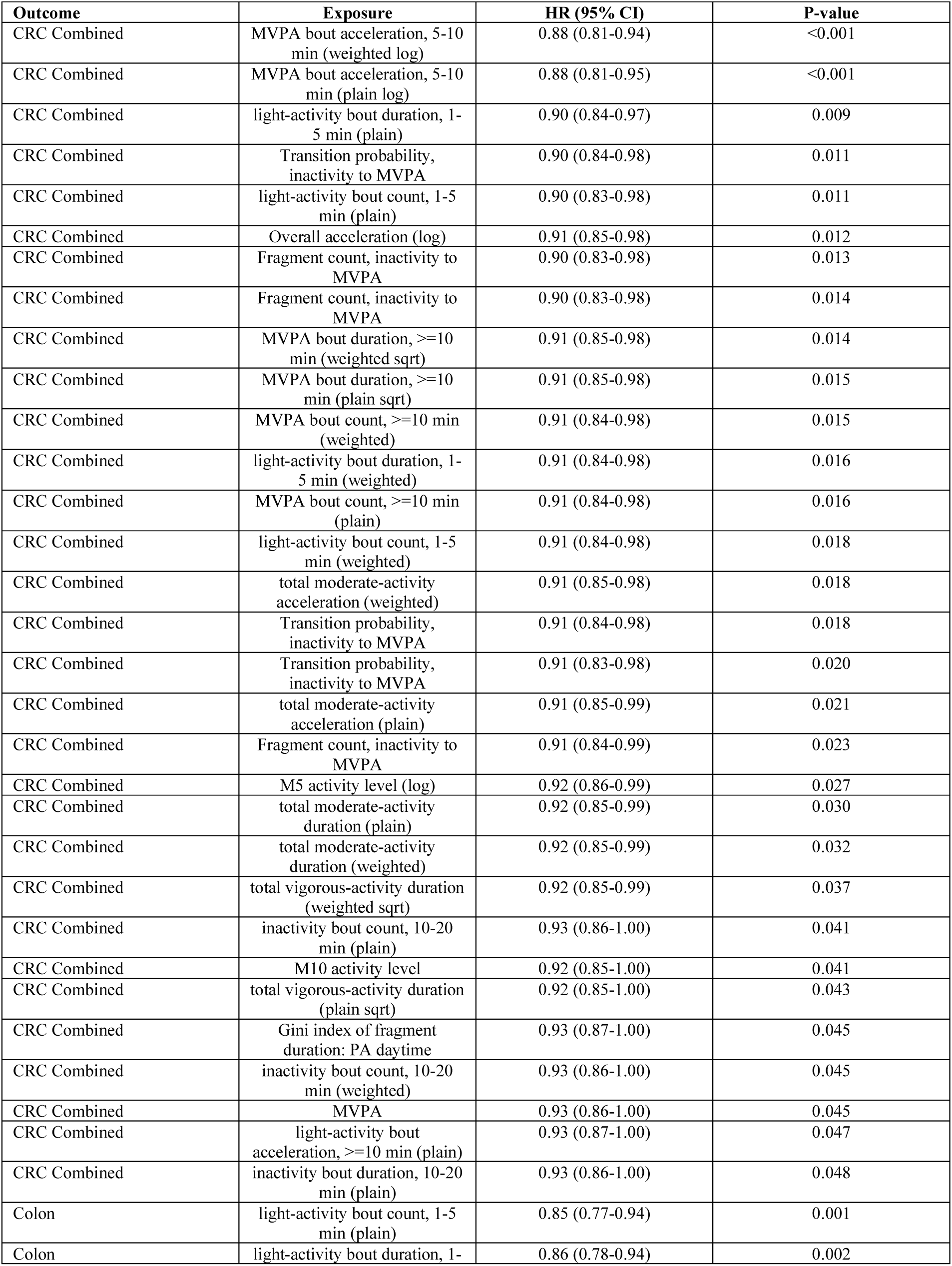
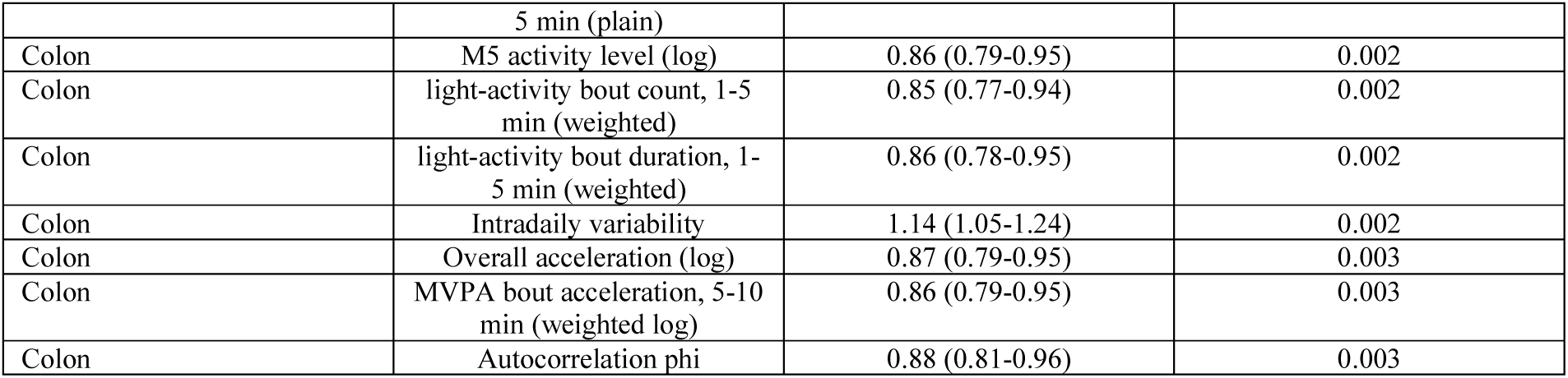
Nominal lead associations between accelerometer-derived metrics and CRC outcomes. Hazard ratios and 95% confidence intervals for all primary-model associations with p<0.05.

### Overall CRC and Colon Cancer

For total CRC, the most prominent associations were identified within the physical activity architecture domain, specifically involving moderate-to-vigorous physical activity (MVPA) bout intensity. Both weighted and standard 5–10-minute MVPA bouts were inversely associated with CRC risk (HR 0.88, 95% CI 0.81–0.94, p = 0.0006; and HR 0.88, 95% CI 0.81–0.95, p = 0.0007, respectively). Additional nominal signals were observed for short-duration light-activity bouts and transitions into MVPA. Colon cancer accounted for the highest signal density in the ExWAS. The strongest signals involved frequent short light-activity bouts (HR 0.85, 95% CI 0.77–0.94; p = 0.0014). Within the circadian and rhythmicity domains, lower activity during the least-active 5-hour window (L5) was protective (HR 0.87, 95% CI 0.79–0.95, p = 0.0016), whereas higher intradaily variability (IV) which is a marker of hourly rhythm fragmentation, was associated with increased risk (HR 1.14, 95% CI 1.05–1.24, p = 0.0024). These findings characterize a broad but heterogeneous behavioral landscape for colon cancer.

### Rectal cancer

In contrast, rectal cancer exhibited a markedly sparse signal profile. Only one metric reached nominal significance: the Self-Similarity Parameter (SSP), a DFA-derived measure of long-range signal correlation, which was associated with increased risk (HR 1.14, 95% CI 1.01–1.28, p = 0.034). Consistent with overall CRC findings, no metrics from the sleep or light domains were associated with rectal cancer risk.

### Dependence on adjustment tier

The nominal signal burden was highly sensitive to model specification. For colon cancer, significant associations decreased from 89 in the Minimal model to 9 in the Independent Pattern model, suggesting that many structural behavioral signals are intrinsically linked to total acceleration volume. Conversely, lag-time analyses demonstrated relative temporal stability; of the 121 primary associations, 99 remained significant after excluding the first two years of follow-up (Table 3), arguing against a purely monotonic collapse driven by reverse causation. E-values for the lead set (ranging from 1.35 to 1.63) suggested that while the associations are notable, they could be explained by moderate unmeasured confounding. No significant interactions by sex or age were identified.

**Table 3.**
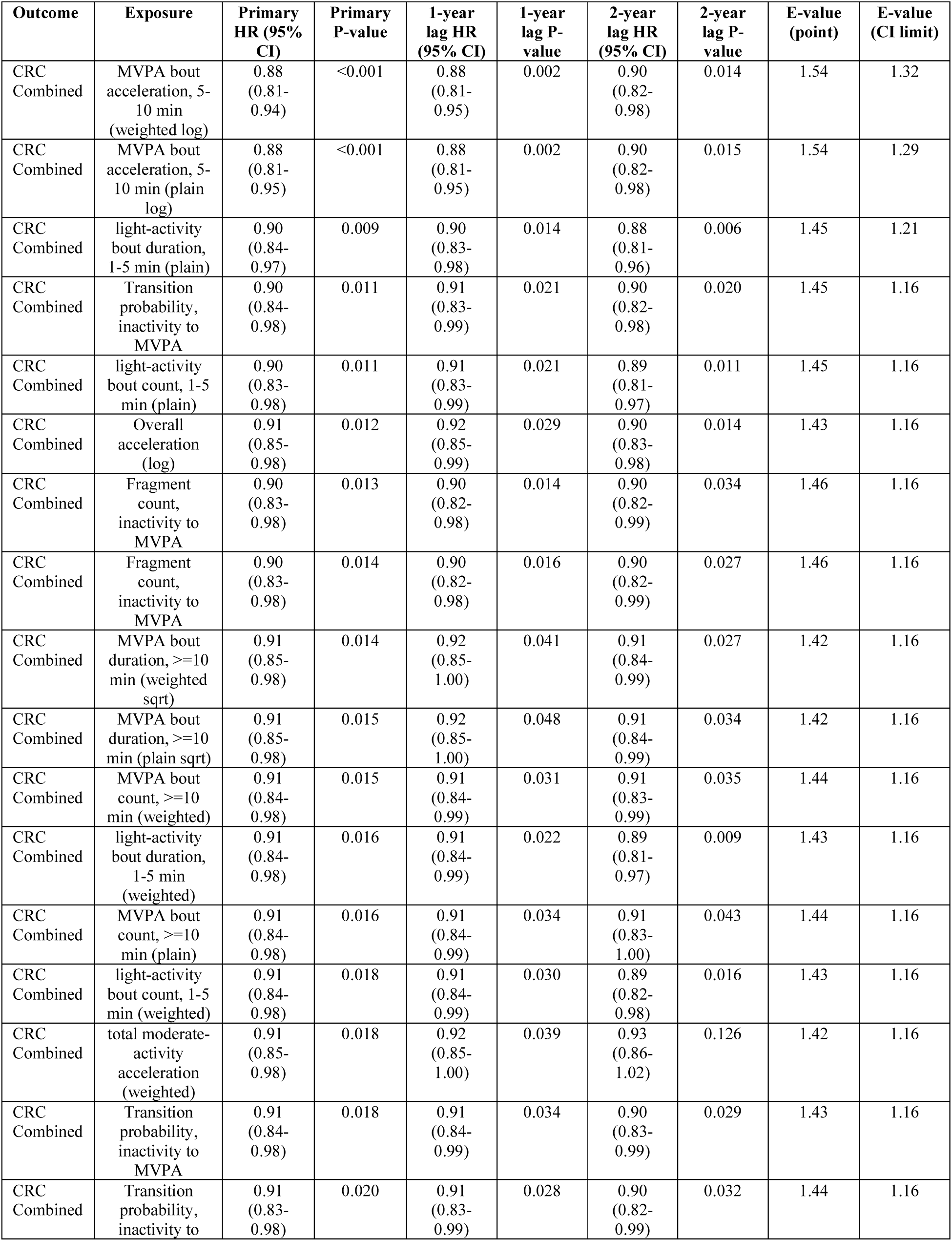
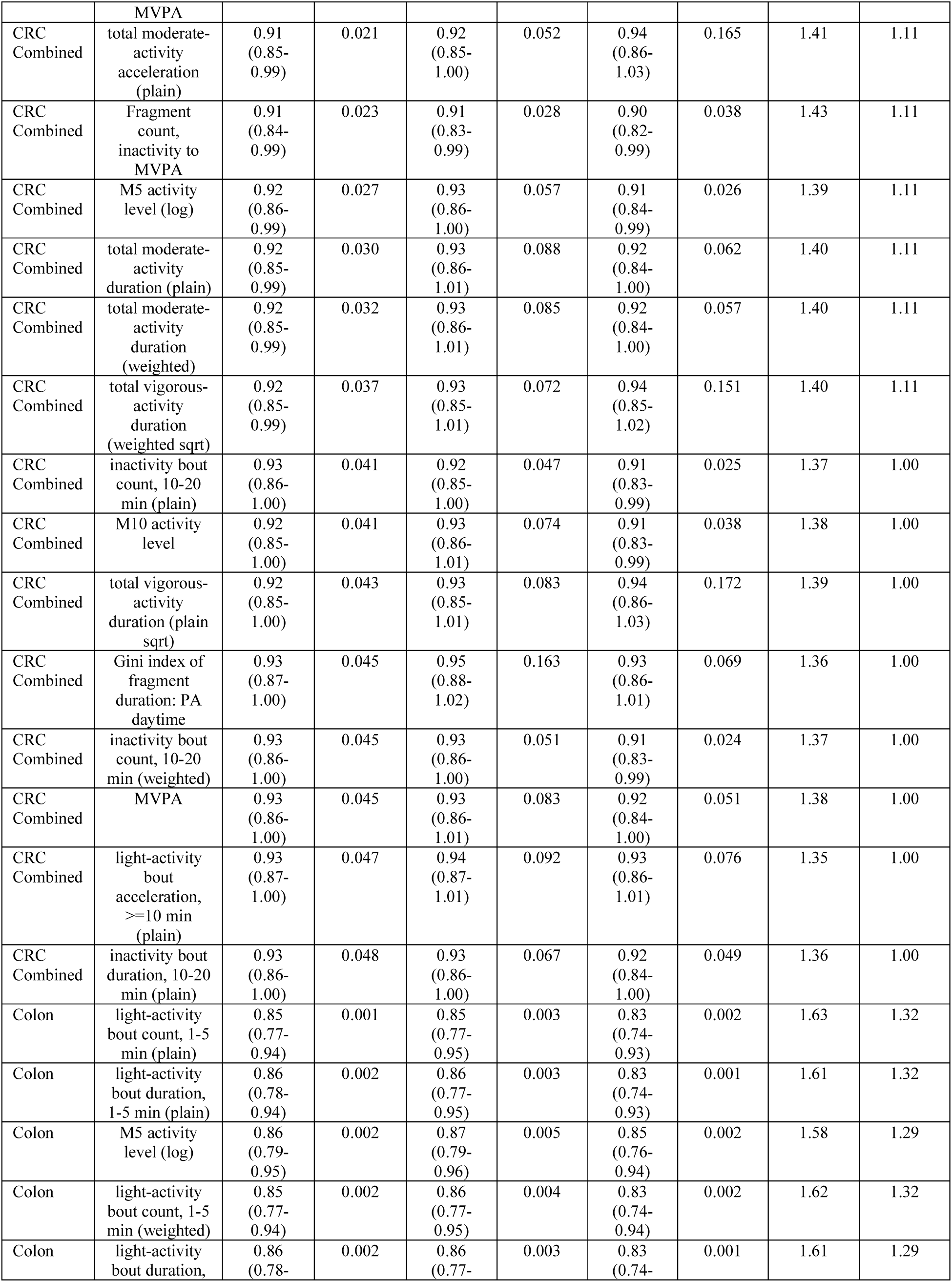
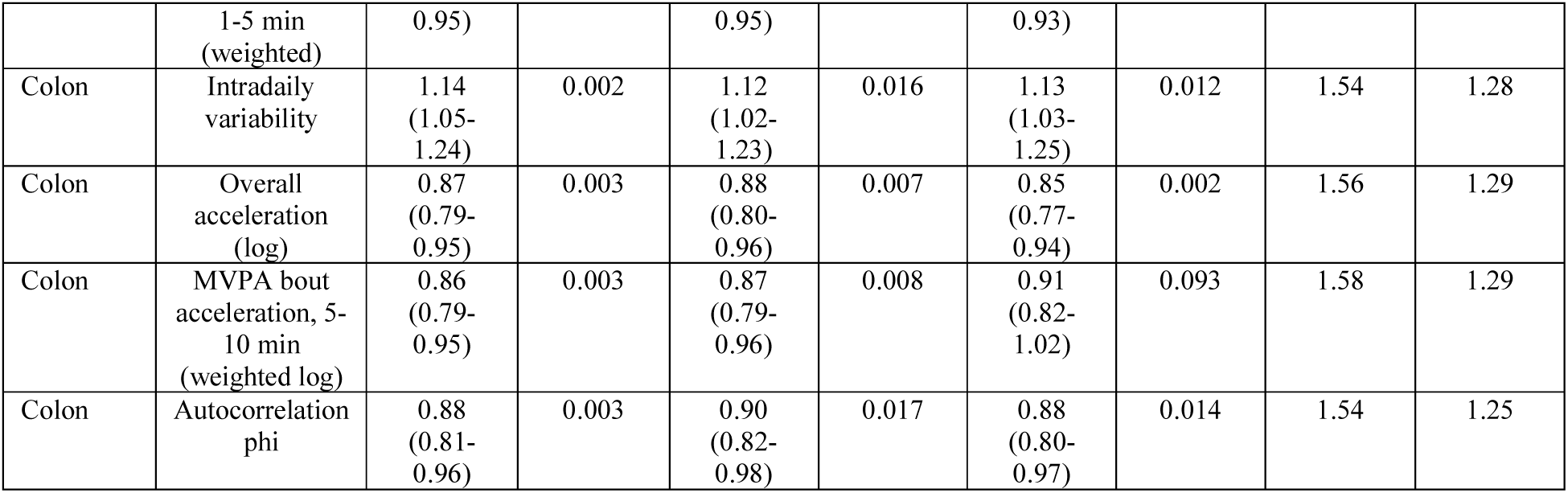
Lag sensitivity analysis results. Comparison of primary, 1-year lag and 2-year lag estimates for the nominal lead set, with corresponding E-values.

### Integrated behavioural profile analysis

To synthesize the five behavioral domains which are light exposure, step counts, physical activity, sleep, and circadian rhythm, we identified three interpretable digital phenotypes through cluster analysis: “high-activity/robust-rhythm,” “low-activity/fragmented-rhythm,” and an intermediate profile. While associations were null for overall CRC and rectal cancer, the “low-activity/fragmented-rhythm” profile was associated with a significantly higher risk of colon cancer (HR 1.33, 95% CI 1.06–1.67; p = 0.015) compared to the intermediate group. However, this multivariate signal showed limited robustness to alternative feature selection, mirroring the sensitivity observed in single-metric models. (Supplementary Tables S9-S10). Moreover, we added baseline blood biochemistry measurements, self-reported chronotype questionnaire and proteomics/metabolomics data into the cluster analysis however the three interpretable digital phenotypes remain the same.

## Discussion

In this hypothesis-generating exposure-wide association study (ExWAS) of 95,050 UK Biobank participants, we leveraged high-resolution accelerometry to define a multidimensional circadian framework encompassing light exposure, step counts, sleep architecture, physical activity, and circadian rhythmicity. Our analysis identified a broad landscape of nominal associations between these digital phenotypes and colorectal cancer (CRC) risk, particularly for colon cancer. The signal was predominantly concentrated within the physical activity architecture and locomotor step count domains, whereas metrics for nocturnal sleep and environmental light exposure did not reach nominal significance. These findings suggest that in this large-scale cohort, the behavioral signature of CRC risk is carried primarily by daytime movement structure and rhythm fragmentation rather than by isolated sleep or light parameters^38, 39^.

Traditional epidemiologic studies of CRC have long relied on self-reported physical activity, which often fails to capture the temporal nuances of human behavior. By integrating five distinct behavioral domains, our study provides a granular map of how the 24-hour cycle relates to oncogenesis. The prominence of step-count metrics (specifically peak cadence) and physical activity architecture (short-bout intensity) adds a layer of structural detail previously inaccessible^22^. This granularity is biologically salient; the concentration of signals around frequent, high-intensity bouts and rhythmic step loading aligns with evidence that sedentary-time interruptions improve metabolic profiles and systemic inflammation^40^, both critical drivers of colorectal carcinogenesis.

A key contribution of this work is the characterization of circadian rhythmicity using both parametric and non-parametric metrics. For colon cancer, the association with higher intradaily variability (IV) which is a marker of rhythm fragmentation^25^ and lower relative amplitude (RA) ^28^suggests that a weakened contrast between daytime activity and nocturnal rest may be an early digital biomarker of risk. This is consistent with preclinical models showing that circadian clock disruption accelerates intestinal tumorigenesis via Apc loss of heterozygosity and altered stem-cell signaling^41–43^. Notably, we observed a marked divergence between subsites: colon cancer exhibited a dense signal burden across multiple domains, while rectal cancer yielded only a single nominal hit in the Self-Similarity Parameter (SSP)^25^. While statistical power may play a role, this divergence likely reflects the distinct anatomical and molecular characteristics of the colon and rectum^10, 44^, suggesting that "colorectal" cancer should perhaps not be viewed as a monolithic behavioral entity^45^.

The lack of nominal associations within the light exposure and sleep architecture domains is informative. While light is the primary zeitgeber^14^ for the master clock, and sleep is essential for immune homeostasis, their individual contributions to CRC risk in this cohort appear less pronounced than the daytime movement phenotype^45^. This may indicate that the "active" phase of the circadian cycle which quantified here by step counts and activity bouts, serves as a more robust integrated marker of circadian health than ambient light or sleep duration alone^39, 46^. However, we cannot exclude the possibility that light-driven effects are more non-linear or that current wrist-worn sensors lack the sensitivity to isolate subtle light-oncogenesis pathways in an older adult population.^19^

Our study underscores the complexity of interpreting multidimensional digital biomarkers^39^. The substantial attenuation of signals after adjusting for adiposity and metabolic factors indicates that much of the circadian behavioral phenotype is intertwined with overall metabolic health. Furthermore, the sensitivity of pattern-based metrics to adjustment for total acceleration suggests that the rhythmic structure of behavior and its total volume are highly correlated, making it difficult to isolate independent causal pathways^6^. Nevertheless, the temporal stability observed in our lag-time analyses, where many associations persisted after excluding early follow-up years, argues against a simple reverse-causation explanation.

The primary strength of this study lies in its objective, multidimensional definition of circadian behavior in a massive prospective cohort. By implementing a transparent ExWAS across five domains, we have mapped a candidate behavioral landscape for CRC that moves beyond total activity volume. However, several limitations remain. As a hypothesis-generating screen of 224 correlated exposures, our findings require independent replication. Accelerometry was measured at a single time point, and residual confounding from diet or screening behavior is possible. Finally, the “healthy volunteer” bias of the UK Biobank may limit generalizability to higher-risk populations^47^.

## Conclusion

In summary, our five-domain circadian phenotyping reveals that locomotor step counts, physical activity intensity, and circadian rhythmicity are the most promising candidate biomarkers for colon cancer risk. The absence of independent signals for sleep and light exposure suggests that daytime behavioral organization may be the primary driver of the observed associations. Rather than providing a definitive list of risk factors, this study provides a systematic map to prioritize specific behavioral domains for future mechanistic research and clinical replication. These results emphasize that the temporal structure of behavior, captured through the lens of circadian biology, is a critical dimension of cancer epidemiology.

**Figure 1.**
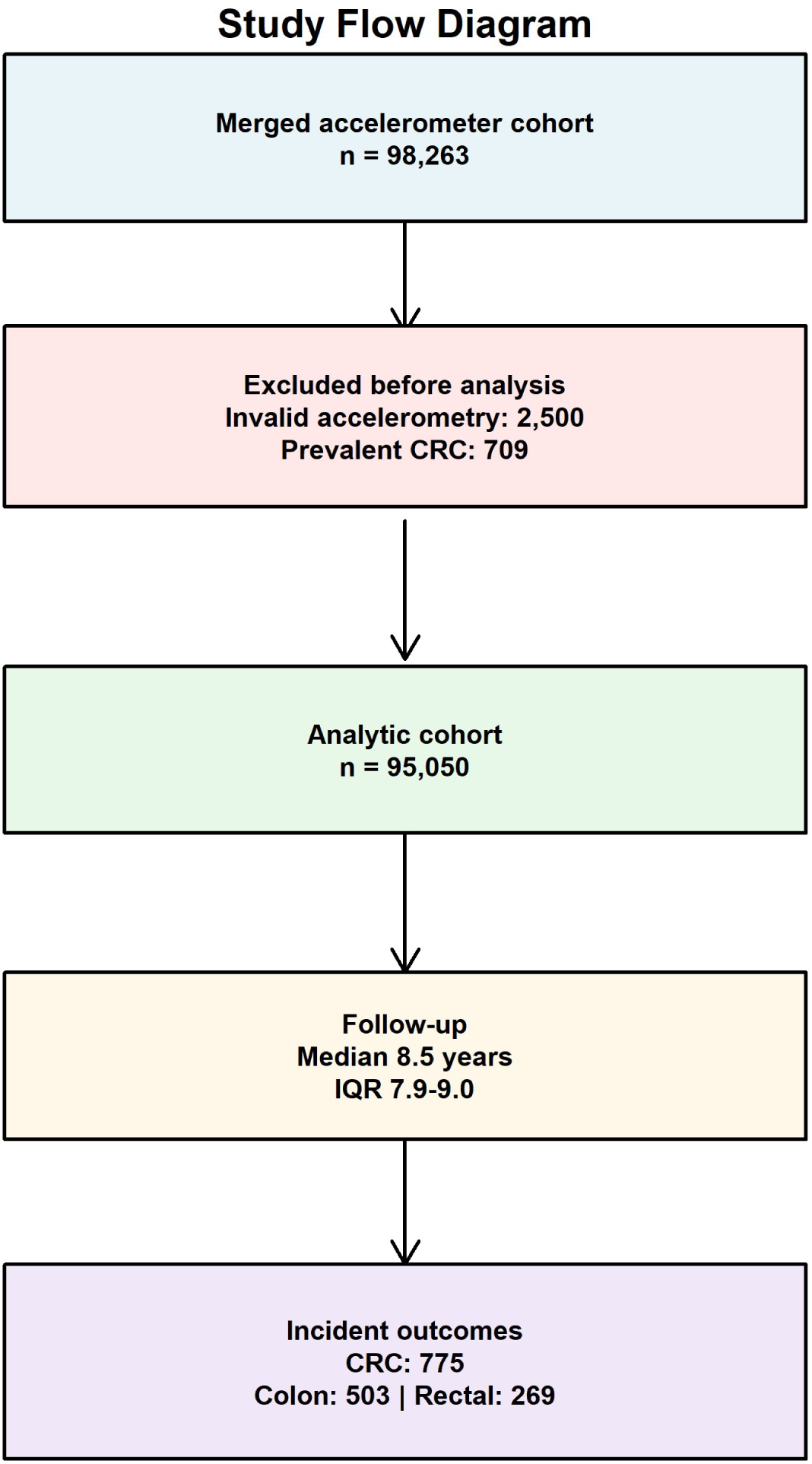
Study flow diagram. Participant flow from the merged accelerometer cohort to the analytic sample and incident CRC outcomes.

**Figure 2.**
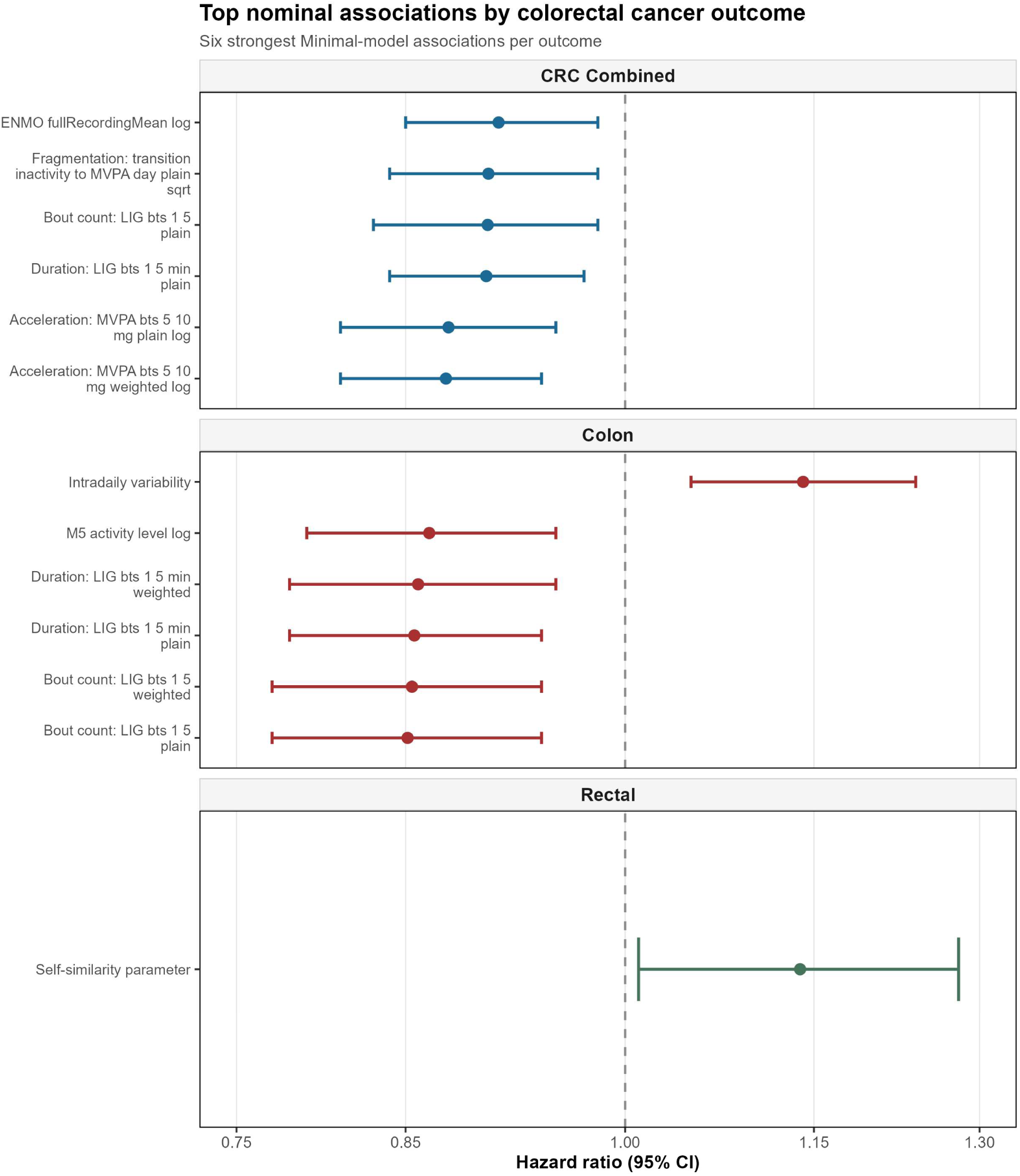
Forest plot of top nominal associations. Forest plot showing the six strongest Minimal-model associations per outcome, stratified by CRC combined, colon cancer and rectal cancer.

**Figure 3.**
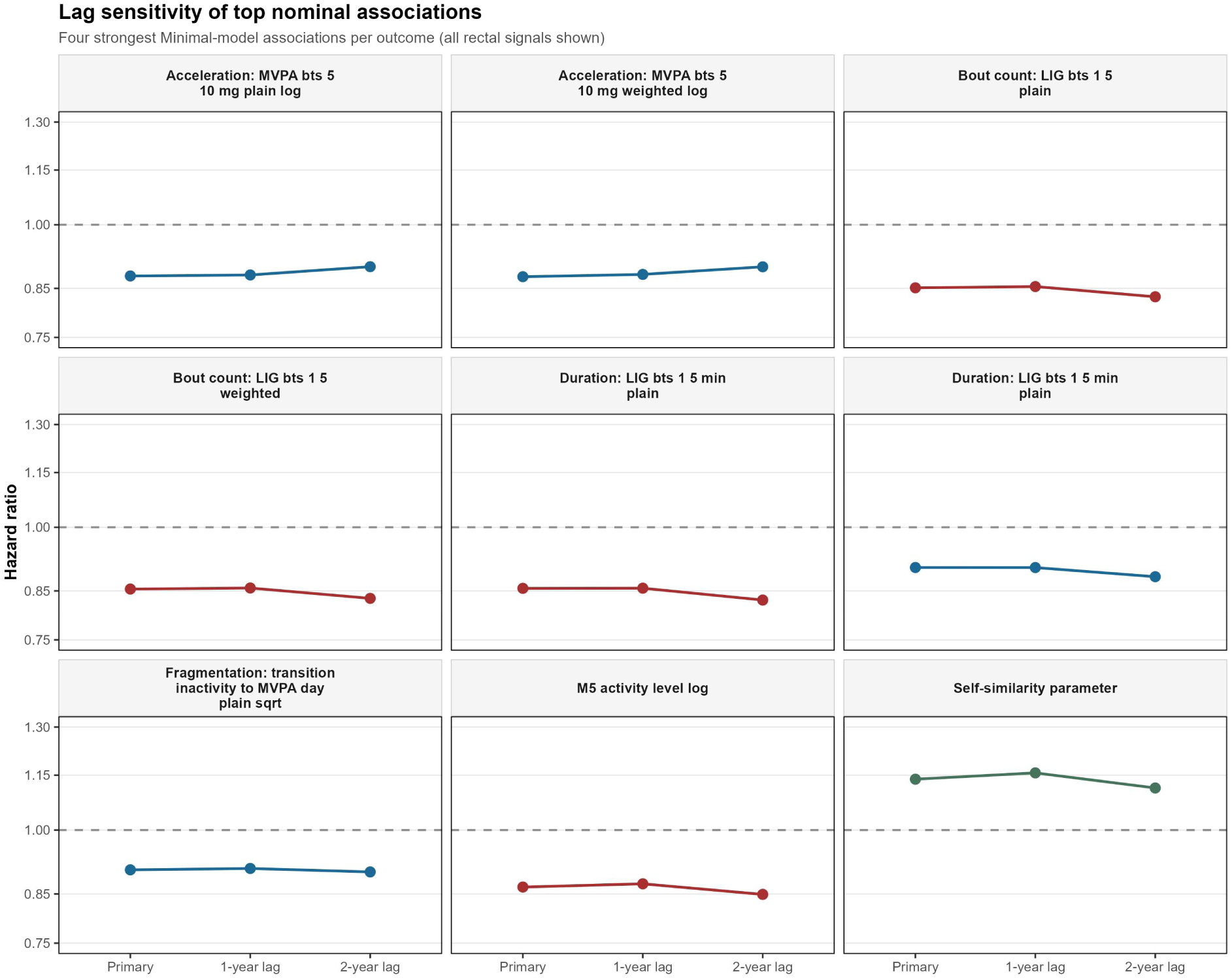
Lag sensitivity analyses. Hazard ratios for the strongest nominal associations across the primary, 1-year lag and 2-year lag models.

## Supporting information

Supplementary Table 2. Full primary-model results for colon cancer

Supplementary Table 3. Full primary-model results for rectal cancer

Supplementary Table 4. Full tiered-model results for CRC combined

Supplementary Table 5. Full tiered-model results for colon cance

Supplementary Table 6. Full tiered-model results for rectal cancer

Supplementary Table 7. Proportional hazards tests for nominal lead associations

Supplementary Table 8. E-values for the nominal lead set

Supplementary Table 9. Integrated behavioural cluster profiles spanning light, step count, physical activity, sleep and circadian rhythm domains

Supplementary Table 10. Associations between integrated behavioural clusters and CRC outcomes

Supplementary Table 1. Full primary-model results for CRC combined.

## Data Availability

All data are available online at https://www.ukbiobank.ac.uk/.

## Ethical approval

The North West Multi-Centre Research Ethics Committee approved the protocol of UK Biobank (REC reference: 11/NW/0382), and all participants provided informed consent before data collection

## Data Sharing

UK Biobank data are available at https://www.ukbiobank.ac.uk/.

## Author Contributions

J.A.B. designed the study, performed statistical analyses.

M.N.C.C. interpreted results and wrote the manuscript.

## Supplementary Information

**Supplementary Table 1.** Full primary-model results for CRC combined.

**Supplementary Table 2.** Full primary-model results for colon cancer.

**Supplementary Table 3.** Full primary-model results for rectal cancer.

**Supplementary Table 4.** Full tiered-model results for CRC combined.

**Supplementary Table 5.** Full tiered-model results for colon cancer.

**Supplementary Table 6.** Full tiered-model results for rectal cancer.

**Supplementary Table 7.** Proportional hazards tests for nominal lead associations.

**Supplementary Table 8.** E-values for the nominal lead set.

**Supplementary Table 9.** Integrated behavioural cluster profiles spanning light, step count, physical activity, sleep and circadian rhythm domains.

**Supplementary Table 10.** Associations between integrated behavioural clusters and CRC outcomes.

